# Effectiveness of Plyometric Exercises Versus Flatfoot Corrective Exercises on Balance and Foot Posture in Obese Children with a Flexible Flatfoot: A Comparative Study

**DOI:** 10.64898/2026.10.31.26361899

**Authors:** Mahendrakar Manimala, Pallavi wajapey

## Abstract

**Background and Purpose:** Childhood obesity often leads to flexible flatfoot, which negatively affects foot posture, balance, and physical function. While exercise is commonly used in clinical practice, there is limited comparative evidence on plyometric training versus traditional corrective exercises in this population. This study aimed to compare the effects of an 8-week plyometric exercise program versus flatfoot corrective exercises on balance and foot posture in obese children aged 7–11 years with flexible flatfoot.

**Methods:** A parallel, non-randomized comparative study was conducted with 120 children recruited from schools in Bengaluru. Participants were allocated to either Group A (plyometric exercises, n = 60) or Group B (flatfoot corrective exercises, n = 60). Both groups trained for 8 weeks (two 40-minute sessions weekly). Foot posture was assessed using the Foot Posture Index-6 (FPI-6), and static balance was measured as center-of-pressure path length using the BTrackS Balance Tracking System before and after the intervention.

**Results:** Both groups demonstrated statistically significant within-group improvements in foot posture and balance after 8 weeks (p < 0.001). However, the plyometric group achieved slightly better improvements in foot posture (statistically significant for the left foot, p = 0.028) and markedly greater gains in balance (median path length: 30.39 cm vs. 37.45 cm, p< 0.001) compared to the corrective exercise group.

**Conclusions:** Both exercise approaches are beneficial for obese children with flexible flatfoot. Plyometric training provides additional advantages in improving dynamic balance and neuromuscular control, supporting its inclusion in pediatric physiotherapy programs.

## 1. Introduction

Childhood obesity has emerged as a major global public health challenge, currently affecting over 380 million children and adolescents worldwide [1,2]. In India, rapid socioeconomic and lifestyle transitions have driven a steep rise in pediatric excess weight across both urban and rural demographics, with projections indicating that the country will account for approximately 11% of the global childhood obesity burden by 2030 [1]. South Asian children are particularly vulnerable to the adverse metabolic and musculoskeletal consequences of excess adiposity, with estimates from the World Obesity Atlas projecting that over 83 million children in India will be living with overweight or obesity by 2035 [3].

Excess body mass imposes chronic, abnormally high mechanical loads on the developing musculoskeletal system during critical stages of growth and skeletal maturation [4,5]. In children aged 7–11 years, the medial longitudinal arch is undergoing vital ossification and structural stabilization [5,8]. The foot serves as the primary base of support and sensorimotor interface for the kinetic chain, facilitating weight distribution, shock absorption, and postural balance [4]. In children with obesity, excessive and repetitive plantar loading frequently leads to flexible flatfoot (pes planus), which accounts for nearly 95% of pediatric flatfoot presentations [5–10]. Clinically, flexible flatfoot is characterized by collapse of the medial longitudinal arch, hind-foot valgus deviation, and forefoot abduction upon full weight-bearing, which typically resolves during non-weight-bearing or toe-standing [8–10]. This malalignment alters center-of-gravity displacement over the base of support, impairs static and dynamic postural control, increases center-of-pressure sway velocity, and elevates the risk of lower-limb fatigue and falls [4,6,10].

Conservative physiotherapy management routinely employs flatfoot corrective exercises targeting active and passive arch support [11,12]. These traditional protocols focus on isolated strengthening of the intrinsic foot musculature (such as the abductor hallucis and flexor digitorum brevis) and extrinsic stabilizers (primarily the tibialis posterior and anterior), combined with passive triceps surae stretching to relieve Achilles tendon tightness [11–13]. While corrective exercises effectively enhance localized muscle strength and static arch morphology, daily functional tasks and sport activities require rapid, dynamic stabilization under continuous kinetic perturbations.

Plyometric training utilizes the stretch-shortening cycle (SSC), pairing rapid eccentric lengthening with immediate, explosive concentric muscular contractions through jumping, hopping, bounding, and landing drills [14–16]. This dynamic loading stimulates muscle spindle sensitivity, improves Golgi tendon organ regulation, increases lower-extremity joint stiffness, and enhances feed-forward (anticipatory) and feedback postural adjustments [14,16,17]. By actively challenging the entire “Foot Core System”—the dynamic interaction between the passive structural arch, active muscular stabilizers, and neural mechanoreceptors networks—plyometric may offer unique sensorimotor advantages over localized, static exercises [23].

Although both corrective exercises and plyometric training have demonstrated individual benefits in pediatric rehabilitation, direct comparative evidence evaluating their relative efficacy on foot alignment and objective postural sway metrics specifically in obese children remains sparse. Furthermore, clinical investigations addressing this problem within the Indian pediatric population are notably limited.

Therefore, this study was conducted to compare the effects of an 8-week plyometric exercise program versus a conventional flatfoot corrective exercise program on foot posture and balance in obese children aged 7–11 years with flexible flatfoot. We hypothesized (H1) that an 8-week plyometric exercise program would yield significantly greater improvements in static balance and foot posture compared to conventional flatfoot corrective exercises due to enhanced stretch-shortening cycle conditioning and sensorimotor adaptations, tested against the null hypothesis (H0) that there is no statistically significant difference between the two intervention protocols.

## Materials and Methods

### Source of Data and Study Design

The study was conducted among children recruited from MES High School, R.V. Public School, JSS Higher Primary and High School, and R.V. Girls High School, located in and around Jayanagar, Bengaluru. A comparative study with a parallel, non-randomized design was conducted over a period of six months. Children aged 7– 11 years were recruited from the selected schools using purposive sampling.

### Sample Size

The sample size was calculated using Zα/_2_=1.96, Z1-β=1.282, pooled proportion P=0.124,q=0.876, and an absolute precision of 0.10. The alpha error was set at 0.05 and the beta error at 0.20. The calculated sample size was 101 participants. Considering an additional allowance for attrition, the effective sample size was increased to 120 participants, with 60 participants allocated to each group.

### Ethical Clearance

Ethical approval was obtained from the Institutional Ethics Committee of R.V. College of Physiotherapy, Bengaluru. The trial was prospectively registered with the Clinical Trials Registry–India under registration number CTRI/2026/04/108003. Permission to conduct the study was obtained from the respective school authorities. Written informed consent was obtained from the parents or legal guardians of all participants before enrolment.

### Materials

The materials used in the study included an informed consent form, data-collection form, calibrated weighing scale, stadiometer, stainless-steel scale, yoga mat, stepper, BTrackS Balance System, stationery, and first-aid kit.

### Participants

Children aged 7–11 years were included if their parents or legal guardians provided written informed consent, if they voluntarily agreed to participate, and if they had a BMI of 25.00 kg/m^2^ or higher, flexible flatfoot, and balance impairment. Children were excluded if they had a previous lower-limb injury, lower-limb deformity, a history of neurological or other musculoskeletal conditions, visual or auditory impairment, cardiovascular or respiratory conditions, or participation in an obesity-treatment programme during the preceding three months.

A total of 140 children were assessed for eligibility. Twenty children were excluded, of whom 10 did not meet the inclusion criteria and 10 declined to participate. The remaining 120 children were allocated into Group A, which received plyometric exercises, and Group B, which received flatfoot corrective exercises, with 60 participants in each group. Post-intervention assessment was completed for all participants, and 60 participants from each group were included in the final analysis.

### Screening Procedures

Body Mass Index was used to assess body-weight status and was calculated using the formula: BMI = weight in kilograms divided by height in meters squared. Body weight was measured using a calibrated weighing scale, and height was measured using a stadiometer. Measurements were taken with the child standing barefoot, looking straight ahead, and wearing light clothing. BMI was interpreted using the Revised Indian Academy of Pediatrics 2015 growth charts.

The Navicular Drop Test, originally described by Brody, was used to assess the height of the medial longitudinal arch and the degree of foot pronation. The navicular tuberosity was marked, and its height was measured first in a non-weight-bearing position with the subtalar joint in neutral and then during relaxed weight-bearing standing. The difference between the two measurements was recorded in millimeters as the navicular drop. A navicular drop greater than 9 mm was considered indicative of a pronated foot, 5–9 mm was considered neutral, and less than 5 mm was considered indicative of a supinated foot.

The Two-Minute Walk Test was used to assess walking endurance and functional exercise capacity. The test was performed in a quiet, straight, and level indoor corridor using a 15-metre walking path. Participants were instructed to walk continuously back and forth along the pathway at a self-selected comfortable speed for two minutes. The total distance covered was calculated by adding the distance completed during each full lap and the final partial lap and was recorded in meters.

### Outcome Measures

Balance was assessed using the BTrackS Balance Tracking System. The child stood barefoot on the balance plate with the feet positioned shoulder-width apart and the hands resting on the hips. Four trials of 20 seconds each were performed, with the first trial serving as a practice trial and the subsequent three trials being recorded for analysis. Participants were instructed to stand as still as possible with their eyes closed. The system recorded the centre-of-pressure path length in centimeters and other sway parameters. Testing was conducted in a quiet environment with minimal distractions after a brief familiarization session.

Foot posture was assessed using the Foot Posture Index-6. The child stood barefoot in a relaxed bilateral stance on a firm, level surface, with the arms relaxed by the sides and the eyes directed forwards. The six criteria assessed were talar head palpation, curvature of the supra-and infra-lateral malleoli, frontal-plane position of the calcaneus, prominence of the talonavicular joint region, congruence of the medial longitudinal arch, and abduction or adduction of the forefoot in relation to the rear foot. Each item was scored from –2 to +2, giving a total score ranging from –12 to +12. Negative scores indicated a more supinated foot posture, whereas positive scores indicated a more pronated foot posture. Both feet were assessed separately.

## Procedure

Following ethical approval and permission from the school authorities, demographic data were collected and the children were screened using BMI and the Navicular Drop Test. Parents or legal guardians of eligible children were provided with information regarding the purpose and procedures of the study, and written informed consent was obtained. Baseline assessment included evaluation of foot posture using the Foot Posture Index-6 and balance using the BTrackS Balance System. Participants were then allocated into two experimental groups. The respective exercise programme were explained verbally and demonstrated by the physiotherapist before commencement.

### Exercise Programme

For Group A, the plyometric exercise programme was administered for 8 weeks, with two sessions per week, each lasting 40 minutes. Each session included a 5-minute warm-up, the prescribed plyometric exercises, and a 5-minute cool-down consisting of gentle stretching of the lower-limb muscles. The programme included low-intensity lateral jumps, moderate-intensity squat jumps, increasing-height jumps, decreasing-height jumps, jumps involving different platform heights, and single-leg vertical jumps. The exercises were performed on a sponge mat for shock absorption. Participants were instructed to land on their toes, maintain appropriate lower-limb alignment, and control excessive foot pronation during weight-bearing activities.During weeks 1–2, participants performed lateral, squat, different-height, increasing-height, and decreasing-height jumps at 10×5, weeks 3–4 included increasing-height plus squat jumps at 12×5, weeks 5–6 added right single-leg jumps at 14×5, and weeks 7–8 included left and right single-leg jumps at 16×5

For Group B, the flatfoot corrective exercise programme was administered for 8 weeks, with two sessions per week, each lasting 40 minutes. Each session began with a 5-minute warm-up consisting of gentle stretching of the lower-limb muscles. The programme included passive range-of-motion exercises for the ankle and foot, calf-muscle stretching, strengthening of the tibialis anterior, tibialis posterior, flexor hallucis longus, intrinsic foot muscles and plantar muscles, towel-curl exercises, heel raises with a ball, unilateral weight-bearing, toe walking, heel walking, proprioceptive activities, and balance exercises.

Foot posture and balance were assessed before the intervention and reassessed after completion of the respective exercise programme. The same equipment, assessment procedures, and testing conditions were used for both assessments.

## Statistical Analysis

The data were entered and analysed using IBM SPSS Statistics software. Categorical variables were presented as frequencies and percentages, while continuous variables were expressed as mean and standard deviation or median and interquartile range, depending on the distribution of the data. The Shapiro–Wilk test was used to assess normality. Within-group pre-and post-intervention differences were analysed using the paired t-test or Wilcoxon signed-rank test, as appropriate. Between-group comparisons were performed using the independent t-test or Mann–Whitney UUU test. A p-value of ≤0.05 was considered statistically significant.

## Results

### Participant Characteristics

A total of 120 children were included in the analysis. Of these, 64 (53.3%) were boys and 56 (46.7%) were girls. Sixty-six participants (55.0%) were aged 7–9 years and 54 (45.0%) were aged 10–11 years. Sixty participants were allocated to the plyometric exercise group (Group A), and 60 were allocated to the flatfoot corrective exercise group (Group B). The participant characteristics are presented in Table 1.

**Table 1.** Within-Group Comparison of Right-Foot FPI-6 Scores.

| Group | Pre-test median (IQR) | Post-test median (IQR) | Wilcoxon Z-value | p-value |
|---|---|---|---|---|
| Group A: Plyometric exercise | 8.00 (2.00) | 4.00 (2.00) | 6.769 | <0.001 |
| Group B: Flatfoot corrective exercise | 8.00 (2.00) | 5.00 (2.75) | 6.829 | <0.001 |

Statistically significant difference from baseline (p < 0.05).
FPI-6: Foot Posture Index-6; IQR: interquartile range.

### Distribution of Data

The Shapiro–Wilk test demonstrated a non-normal distribution for age, BMI, and all FPI-6 measurements in both groups. The pre-intervention BTrackS path length was normally distributed in Group A but not in Group B, whereas the post-intervention BTrackS path length was normally distributed in both groups. Since most study variables were non-normally distributed, non-parametric tests were used for the analysis. Results are presented as median and interquartile range.

### Foot Posture Index-6

At baseline, the median right-foot FPI-6 score was 8.00 in both groups, while the median left-foot score was 8.00 in Group A and 7.00 in Group B. Following the intervention, the median right-foot FPI-6 score decreased to 4.00 in Group A and 5.00 in Group B. The median left-foot score decreased to 4.00 in both groups.

Within-group analysis demonstrated a statistically significant reduction in right-and left-foot FPI-6 scores in both the plyometric exercise group and the flatfoot corrective exercise group (p<0.001). Between-group analysis showed no statistically significant difference in the right-foot (p=0.703)or left-foot (p=0.464) FPI-6 scores at baseline. Following the intervention, the between-group difference was not statistically significant for the right foot (p=0.077); however, a statistically significant difference was observed for the left foot (p=0.028). The FPI-6 results are presented in Table 2 and Figure 2.

**Table 2.** Between-Group Comparison of Right-Foot FPI-6 Scores.

| **Time point** | **Group A median (IQR)** | **Group B median (IQR)** | **Mann–Whitney Z-value** | **p-value** |
|---|---|---|---|---|
| Pre-test | 8.00 (2.00) | 8.00 (2.00) | 0.703 | 0.480 |
| Post-test | 4.00 (2.00) | 5.00 (2.75) | 1.770 | 0.077 |

Statistically significant difference from baseline (p < 0.05).
FPI-6: Foot Posture Index-6; IQR: interquartile range.

**Table 3.** Within-Group Comparison of Left-Foot FPI-6 Scores.

| **Group** | **Pre-test median (IQR)** | **Post-test median (IQR)** | **Wilcoxon Z-value** | **p-value** |
|---|---|---|---|---|
| Group A: Plyometric exercise | 8.00 (2.00) | 4.00 (2.00) | 6.764 | <0.001 |
| Group B: Flatfoot corrective exercise | 7.00 (3.00) | 4.00 (3.00) | 6.798 | <0.001 |

Statistically significant difference from baseline (p < 0.05).
FPI-6: Foot Posture Index-6; IQR: interquartile range.

**Table 4.** Between-Group Comparison of Left-Foot FPI-6 Scores.

| **Time point** | **Group A median (IQR)** | **Group B median (IQR)** | **Mann–Whitney Z-value** | **p-value** |
|---|---|---|---|---|
| Pre-test | 8.00 (2.00) | 7.00 (3.00) | 0.464 | 0.640 |
| Post-test | 4.00 (2.00) | 4.00 (3.00) | 2.199 | 0.028 |

Statistically significant difference from baseline (p < 0.05).
FPI-6: Foot Posture Index-6; IQR: interquartile range.

**Table 5.**
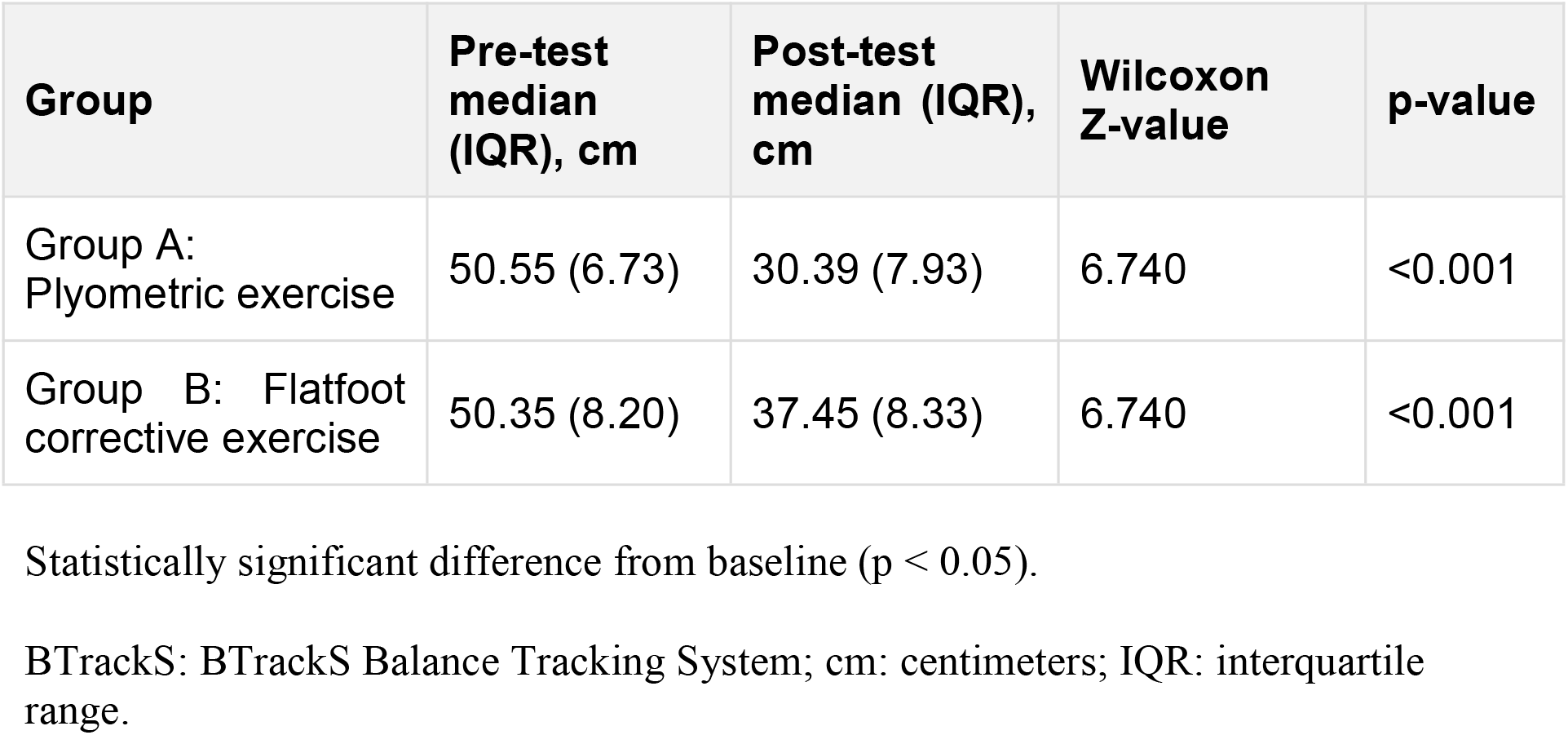
Within-Group Comparison of BTrackS Path Length.

| Group | Pre-test median (IQR), cm | Post-test median (IQR), cm | Wilcoxon Z-value | p-value |
|---|---|---|---|---|
| Group A: Plyometric exercise | 50.55 (6.73) | 30.39 (7.93) | 6.740 | <0.001 |
| Group B: Flatfoot corrective exercise | 50.35 (8.20) | 37.45 (8.33) | 6.740 | <0.001 |

Statistically significant difference from baseline (p < 0.05).
BTrackS: BTrackS Balance Tracking System; cm: centimeters; IQR: interquartile range.

**Table 6.** Between-Group Comparison of BTrackS Path Length.

| **Time point** | **Group A median (IQR), cm** | **Group B median (IQR), cm** | **Mann–Whitney Z-value** | **p-value** |
|---|---|---|---|---|
| Pre-test | 50.55 (6.73) | 50.35 (8.20) | 0.346 | 0.730 |
| Post-test | 30.39 (7.93) | 37.45 (8.33) | 6.099 | <0.001 |

Statistically significant difference from baseline (p < 0.05).
BTrackS: BTrackS Balance Tracking System; cm: centimeters; IQR: interquartile range.

### BTrackS Balance

The median pre-intervention BTrackS path length was 50.55 cm in Group A and 50.35 cm in Group B. After the intervention, the path length decreased to 30.39 cm in Group A and 37.45 cm in Group B, indicating a reduction in postural sway.

Within-group analysis showed a statistically significant reduction in BTrackS path length in both Group A (Z=6.740, p<0.001)and Group B (Z=6.740, p<0.001). The groups did not differ significantly at baseline (p=0.730). However, a statistically significant between-group difference was observed after the intervention (Z=6.099, p<0.001), with a greater reduction in path length observed in the plyometric exercise group.

## Discussion

This study compared plyometric exercises with flatfoot corrective exercises in obese children aged 7–11 years with flexible flatfoot. Foot posture was assessed using the Foot Posture Index-6 (FPI-6), and balance was assessed using the BTrackS Balance System. Both groups demonstrated significant improvements in foot posture and balance after 8 weeks. However, the plyometric group showed slightly greater improvement in foot posture and significantly greater improvement in balance.

Childhood obesity increases plantar loading and may contribute to medial longitudinal arch collapse, heel valgus, excessive pronation, impaired balance, and reduced functional mobility. Previous studies have reported flatter feet, increased pronation, reduced ankle dorsiflexion, and greater foot pain in overweight and obese children.^7,19^ The present findings support the use of exercise-based interventions to address these impairments, particularly in the Indian paediatric population.

The reductions in FPI-6 scores in both groups suggest a shift from excessive pronation towards a more neutral foot posture. These improvements may reflect enhanced weight distribution and active support of the medial longitudinal arch. The benefits observed in the corrective exercise group may be attributed to strengthening of the intrinsic foot muscles, tibialis posterior, and tibialis anterior. Similar improvements in foot posture and arch morphology have been reported following corrective and intrinsic foot-muscle strengthening exercises.^12^

The slightly greater improvement in the plyometric group may be related to the dynamic and weight-bearing nature of jumping, hopping, bounding, and landing activities. Plyometric training challenges the foot and ankle complex, stimulates the stretch–shortening cycle, and enhances neuromuscular coordination and dynamic arch control. Previous studies have also reported improvements in coordination, strength, jumping ability, and motor performance following paediatric plyometric training.^14,15,20^

A significant between-group difference was observed in the left-foot FPI-6 score, whereas the right-foot difference was not significant. This asymmetry may be related to pre-existing differences in foot posture, limb dominance, plantar loading, or neuromuscular control. Bilateral foot asymmetry is common in children; therefore, this finding should be interpreted cautiously and may not indicate a clinically significant imbalance.

The most notable between-group difference was observed in balance performance. Although both interventions reduced postural sway, the improvement was greater following plyometric training. This may be explained by enhanced proprioception, sensorimotor integration, anticipatory postural adjustments, and reactive balance responses during repeated jumping and landing tasks. The BTrackS Balance System was able to detect these improvements in postural stability.

The relationship between improved foot posture and balance may be explained by the role of the foot as the primary interface between the body and the supporting surface. Excessive pronation may alter plantar sensory input and lower-limb alignment, whereas improved foot control may enhance weight distribution and postural stability.

The Foot Core System theory provides a useful framework for these findings by describing foot stability as the interaction of passive, active, and neural subsystems. Plyometric exercises may challenge all three systems, whereas corrective exercises primarily target active muscular support.

The findings are consistent with previous studies reporting positive effects of plyometric and corrective exercises in children with flexible flatfoot and obesity.^6,16,17^ Both interventions may therefore be useful in physiotherapy management. However, plyometric exercises may provide additional benefits for dynamic balance and neuromuscular control. A progressive programme combining corrective exercises with appropriately dosed plyometric training may be beneficial, although this approach requires confirmation through randomized controlled trials.

## Limitations

The intervention duration was limited to 8 weeks, and the long-term sustainability of the improvements remains unknown. Participants were recruited from selected schools in Bengaluru and included only obese children aged 7–11 years with flexible flatfoot, limiting generalizability.

The non-randomized design increases the possibility of selection bias and limits causal interpretation. Daily physical activity, footwear, recreational sports, and home-based exercise were not monitored and may have influenced the results. In addition, no combined-treatment group was included.

Only FPI-6 and BTrackS scores were assessed. Gait, plantar pressure, arch height, lower-limb strength, pain, physical activity, and quality-of-life measures were not evaluated. The sample size also limited subgroup analyses, including sex differences, limb dominance, and bilateral foot asymmetry.

### Recommendations

Future research should:

- Use a three-arm randomized controlled design comparing corrective exercises, plyometric exercises, and combined training.
- Extend the intervention to 12–16 weeks and include 3-, 6-, and 12-month follow-up assessments.
- Recruit larger, multisite samples with broader age and weight ranges.
- Monitor physical activity, footwear, adherence, and recreational exercise.
- Include gait analysis, plantar-pressure assessment, arch-height measurements, strength testing, pain, and quality-of-life outcomes.
- Assess baseline limb dominance, bilateral foot posture, plantar loading, and lower-limb alignment.
- Use progressive, supervised training with appropriate landing mechanics and safety monitoring.

### Clinical Implications

Flatfoot corrective exercises may improve active arch support, while plyometric exercises may provide additional benefits for dynamic balance and neuromuscular control. Plyometric activities should be introduced progressively after children demonstrate adequate strength, balance, and landing technique. Regular reassessment using the FPI-6 and BTrackS may assist clinical monitoring.

## Conclusion

Both plyometric and flatfoot corrective exercises significantly improved foot posture and balance in obese children aged 7–11 years with flexible flatfoot after 8 weeks. Plyometric training produced slightly greater improvements in foot posture and significantly greater gains in balance, possibly due to enhanced proprioception and neuromuscular control. Progressive plyometric exercises may therefore complement traditional corrective exercises in physiotherapy management. However, longer-term randomized studies with larger samples and combined intervention protocols are needed to confirm these findings.

## Data Availability

Data Availability Statement
All data produced in the present study are available upon reasonable request to the authors

https://ctri.nic.in/Clinicaltrials/regtrial.php?modid=1&compid=19&EncHid=89386.37145

## Ethical Approval Statement

Ethical clearance was obtained from the Institutional Ethics Committee (IEC) of RV College of Physiotherapy, Bengaluru, Karnataka, India (Approval Reference Number: RVCP/RESEARCH/2025-26/77) prior to the commencement of the study. The study was conducted in strict accordance with the ethical standards of the Declaration of Helsinki. Permissions from the authorities of all participating schools were secured.

Written informed consent was obtained from parents/legal guardians, and assent was confirmed from all participating children before enrollment.

### Trial Registration

The study was registered with the Clinical Trials Registry - India (CTRI) under registration number **CTRI/2026/04/108003**.

## Abbreviation List

2MWT: 2-Minute Walk Test
BMI: Body Mass Index
BOS: Base of Support
COG: Center of Gravity
CoP: Center of Pressure
CTRI: Clinical Trials Registry - India
FPI-6: Foot Posture Index-6
IAP: Indian Academy of Pediatrics
IEC: Institutional Ethics Committee
IQR: Interquartile Range
NDT: Navicular Drop Test
PROM: Passive Range of Motion
ROM: Range of Motion
SSC: Stretch-Shortening Cycle
WHO: World Health Organization

## Acknowledgement

The authors express their sincere gratitude to the school authorities, principals, physical education teachers, and administrative staff of the participating schools in Bengaluru for granting permission and facilitating the testing and exercise sessions. Sincere appreciation is extended to all the participating children and their parents or legal guardians for their active cooperation, compliance, and dedication throughout the 8-week intervention protocol. The authors also thank the management and Department of Pediatric Physiotherapy at RV College of Physiotherapy for providing institutional facilities and technical infrastructure.

## Patient’s Privacy and Consent for Publication

The manuscript does not contain any individual personal identifiers, facial photographs, or identifiable patient details. Written parental/guardian consent and child assent were obtained for the publication of anonymized aggregate research data.

## Author’s Contributions

**Mahendrakar Manimala:** Conceptualization, Methodology, Investigation, Data Curation, Formal Analysis, Writing – Original Draft.

**Pallavi Wajapey:** Conceptualization, Methodology, Supervision, Project Administration, Validation, Writing – Review & Editing.

## Data Availability Statement

The datasets generated and analyzed during the current study are available from the corresponding author upon reasonable request.

---

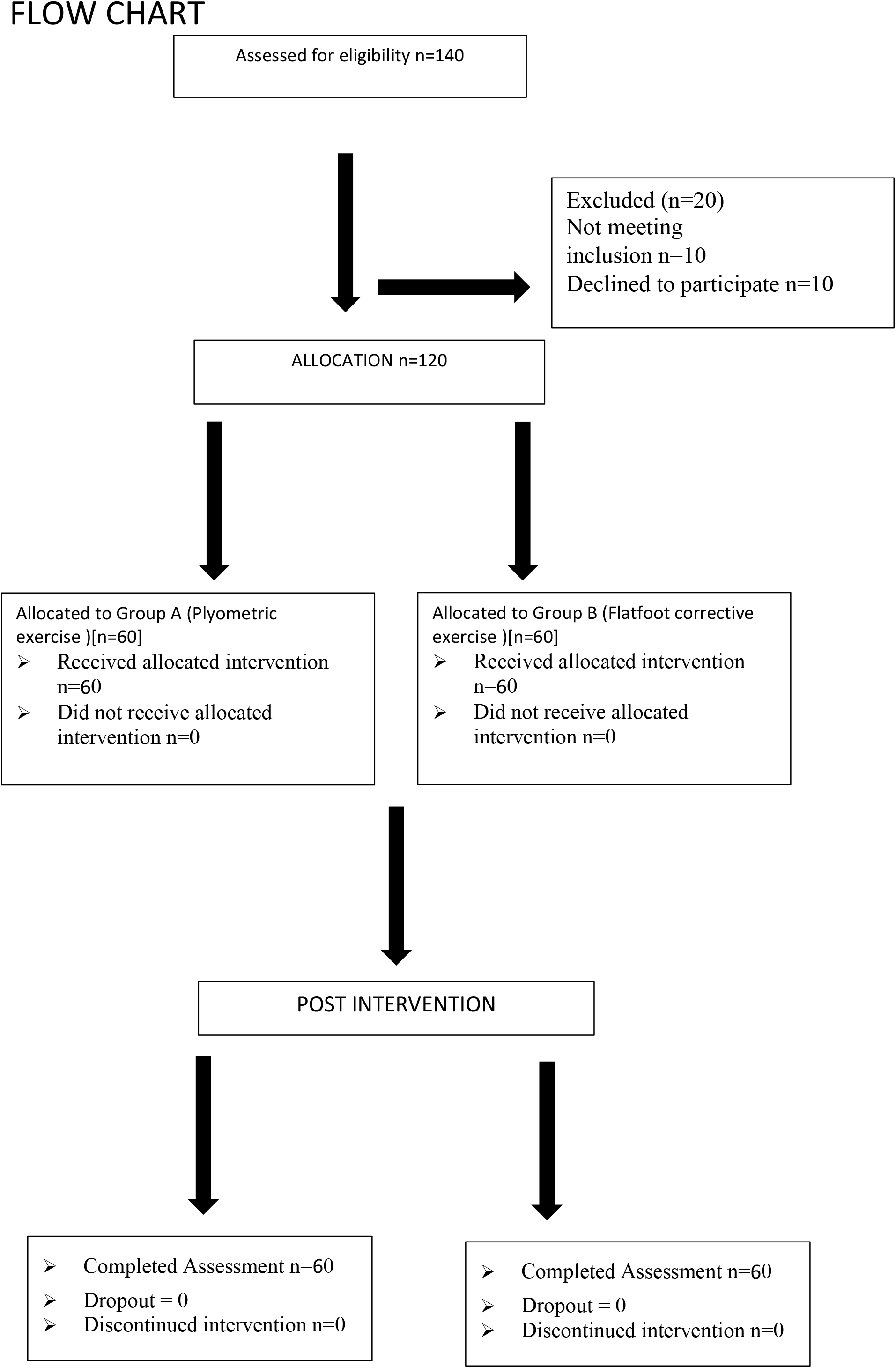
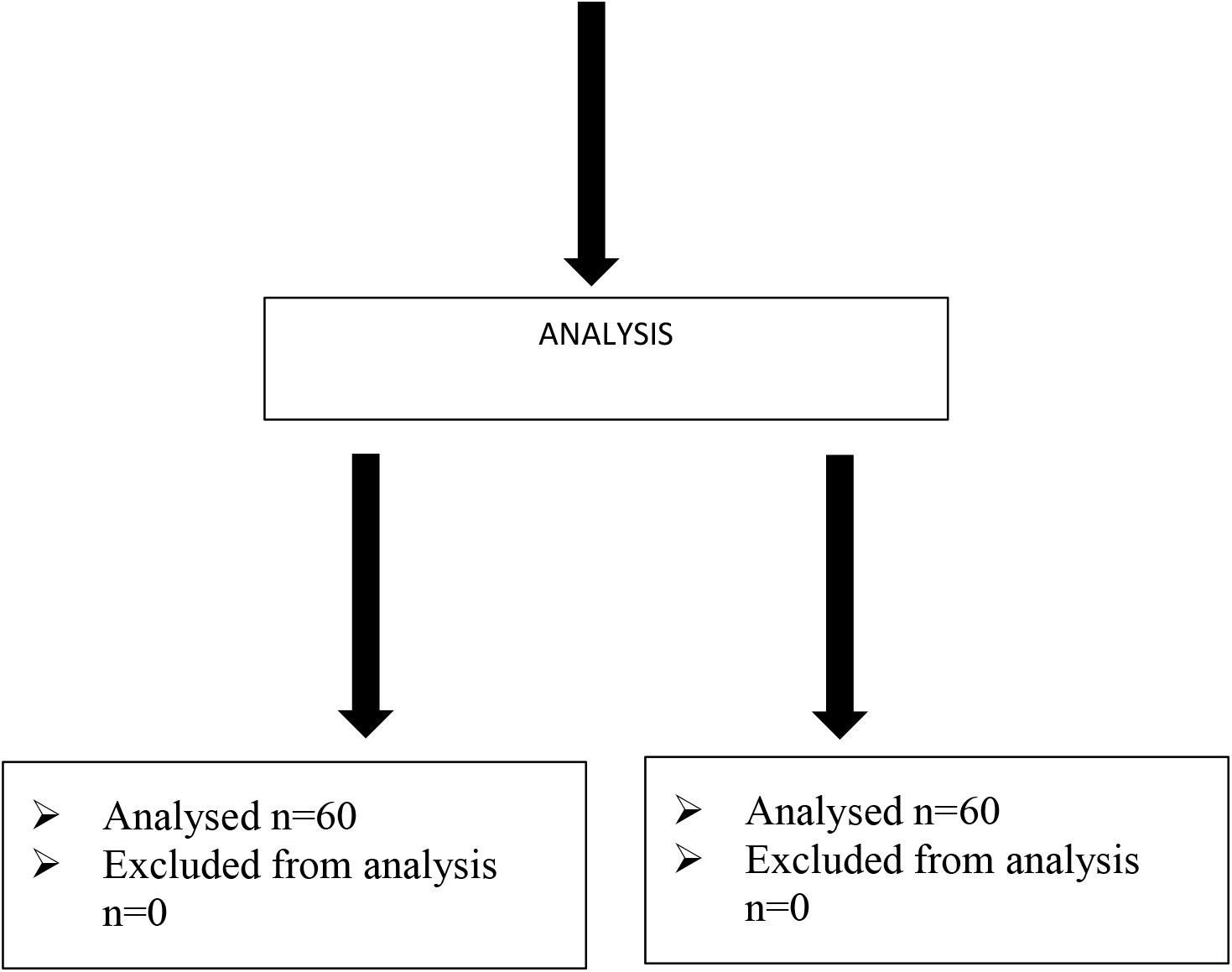

## The Foot Posture Index^©^ FPI-6

### Reference Sheet

The patient should stand in their relaxed stance position with double limb support. The patient should be instructed to stand still, with their arms by the side and looking straight ahead. It may be helpful to ask the patient to take several steps, marching on the spot, prior to settling into a comfortable stance position. During the assessment, it is important to ensure that the patient does not swivel to try to see what is happening for themselves, as this will significantly affect the foot posture. The patient will need to stand still for approximately two minutes in total in order for the assessment to be conducted. The assessor needs to be able to move around the patient during the assessment and to have uninterrupted access to the posterior aspect of the leg and foot.

If an observation cannot be made (e.g. because of soft tissue swelling) simply miss it out and indicate on the datasheet that the item was not scored.

If there is genuine doubt about how high or low to score an item always use the more conservative score.

| **Rearfoot Score** | **-2** | **-1** | **0** | **1** | **2** |
|---|---|---|---|---|---|
| Talar head palpation | Talar head palpable on lateral side/but not on medial side | Talar head palpable on lateral side/slightly palpable on medial side | Talar head equally palpable on lateral and medial side | Talar head slightly palpable on lateral side/ palpable on medial side | Talar head not palpable on lateral side/ but palpable on medial side |
| Curves above and below the malleoli | Curve below the malleolus either straight or convex | Curve below the malleolus concave, but flatter/ more shallow than the curve above the malleolus | Both infra and supra malleolar curves roughly equal | Curve below malleolus more concave than curve above malleolus | Curve below malleolus markedly more concave than curve above malleolus |
| Calcaneal inversion/eversion | More than an estimated 5° inverted (varus) | Between vertical and an estimated 5° inverted (varus) | Vertical | Between vertical and an estimated 5° everted (valgus) | More than an estimated 5° everted (valgus) |
| **Forefoot Score** | **-2** | **-1** | **0** | **1** | **2** |
| Talo-navicular congruence | Area of TNJ markedly concave | Area of TNJ slightly, but definitely concave | Area of TNJ flat | Area of TNJ bulging slightly | Area of TNJ bulging markedly |
| Medial arch height | Arch high and acutely angled towards the posterior end of the medial arch | Arch moderately high and slightly acute posteriorly | Arch height normal and concentrically curved | Arch lowered with some flattening in the central portion | Arch very low with severe flattening in the central portion – arch making ground contact |
| Forefoot abd/adduction | No lateral toes visible. Medial toes clearly visible | Medial toes clearly more visible than lateral | Medial and lateral toes equally visible | Lateral toes clearly more visible than medial | No medial toes visible. Lateral toes clearly visible |

*For further information, manuals and extra datasheets see: www.leeds.ac.uk/medicine/FASTER/FPI/*

## References

1. Singh S, Awasthi S, Kapoor V, Mishra P. Childhood obesity in India: A two-decade meta-analysis of prevalence and socioeconomic correlates. Clin Epidemiol Glob Health. 2023;23:101390. 10.1016/j.cegh.2023.101390

2. World Health Organization. Obesity and overweight. Fact sheet. World Health Organization; 2024.

3. World Obesity Federation. World Obesity Atlas 2024. London: World Obesity Federation; 2024.

4. Alice A, Yadav M, Verma R, Kumari M, Arora S. Effect of obesity on balance: A literature review. Int J Health Sci. 2022;6(S4):3261–3279. 10.53730/ijhs.v6nS4.9192

5. Stolzman S, Irby MB, Callahan AB, Skelton JA. Pes planus and paediatric obesity: a systematic review of the literature. Clin Obes. 2015;5(2):52–59. 10.1111/cob.12089

6. Allam HH, Muhsen A, Al-walah MA, Alotaibi AN, Alotaibi SS, Elsayyad LK. Effects of Plyometric Exercises versus Flatfoot Corrective Exercises on Postural Control and Foot Posture in Obese Children with a Flexible Flatfoot. Appl Bionics Biomech. 2021;2021:3635660. 10.1155/2021/3635660

7. Sadeghi-Demneh E, Azadinia F, Jafarian F, et al. Flatfoot and obesity in school-age children: a cross-sectional study. Clin Obes. 2016;6(1):42–50. 10.1111/cob.12127

8. Živković D, Karaleic S, Andelkovic I. Flat feet and obesity among children. FU Phys Ed Sport. 2018;16(2):347–356. 10.22190/FUPES180602030Z

9. Pfeiffer M, Kotz R, Ledl T, Hauser G, Sluga M. Prevalence of Flat Foot in Preschool-Aged Children. Pediatrics. 2006;118(2):634–639. 10.1542/peds.2005-2126

10. Mosca VS. Flexible flatfoot in children and adolescents. J Child Orthop. 2010;4(2):107–121. 10.1007/s11832-010-0239-9

11. Riccio I, Gimigliano F, Gimigliano R, Porpora G, Iolascon G. Rehabilitative treatment in flexible flatfoot: a perspective cohort study. Musculoskelet Surg. 2009;93(3):101–107. 10.1007/s12306-009-0037-z

12. Carr JB, Yang S, Lather LA. Pediatric Pes Planus: A State-of-the-Art Review. Pediatrics. 2016;137(3):e20151230. 10.1542/peds.2015-1230

13. Halabchi F, Mazaheri R, Mirshahi M, Abbasian L. Pediatric flexible flatfoot; clinical aspects and algorithmic approach. Iran J Pediatr. 2013;23(3):247–260.

14. Nobre GG, De Almeida MB, Nobre IG, et al. Twelve Weeks of Plyometric Training Improves Motor Performance of 7-to 9-Year-Old Boys Who Were Overweight/Obese: A Randomized Controlled Intervention. J Strength Cond Res. 2017;31(8):2091–2099. 10.1519/JSC.0000000000001683

15. Sortwell A, Newton M, Marinho DA, Ferraz R, Perlman D. The Effects of an Eight Week Plyometric-based Program on Motor Performance Skills and Muscular Power in 7-8-Year-Old Primary School Students. IJKSS. 2021;9(4):1–8. 10.7575/aiac.ijkss.v.9n.4p.1

16. Arabmomeni A, Mousavi A. The effects of thera band and water resistance corrective exercises on the correction of flexible flat foot and postural control in boy students with overweight. Pars J Med Sci. 2022;20(3):17–27. 10.52547/jmj.20.3.17

17. Han SJ, Paik JK, Kim M, Kim JS. The effects of 12 weeks of plyometric training on body composition, physical strength, and bone mineral density in obese adolescents. J Exerc Rehabil. 2025;21(5):253–258. 10.12965/jer.2546366.183

18. De Almeida MB, Leandro CG, Queiroz DDR, et al. Plyometric training increases gross motor coordination and associated components of physical fitness in children. Eur J Sport Sci. 2021;21(9):1263–1272. 10.1080/17461391.2020.1829079

19. Hadoush H, Qabook Z, Kanan SF, Almasri NA. Foot arch asymmetry and its correlations with physical fitness level and body mass index among school-aged children. Sci Rep. 2025;15(1):40325. 10.1038/s41598-025-40325-1

20. Liu W, Xu L, Wu H, et al. Bilateral Asymmetries of Plantar Pressure and Foot Balance During Walking, Running, and Turning Gait in Typically Developing Children. Bioengineering. 2025;12(2):151. 10.3390/bioengineering12020151

21. Goble DJ, Rauh MJ, Baweja HS. Normative Data for the BTrackS Balance Test Concussion-Management Tool: Results From 10 045 Athletes Aged 8 to 21 Years. J Athl Train. 2019;54(4):439–444. 10.4085/1062-6050-482-17

22. Goble DJ, Baweja HS. Normative Data for the BTrackS Balance Test of Postural Sway: Results from 16,357 Community-Dwelling Individuals Who Were 5 to 100 Years Old. Phys Ther. 2018;98(9):779–785. 10.1093/ptj/pzy068

23. McKeon PO, Fourchet F. Freeing the Foot. Clin Sports Med. 2015;34(2):347–361. 10.1016/j.csm.2014.12.002

